# Seroprevalence of SARS-CoV-2 Antibodies among vaccinated and non-vaccinated adults in the West Bank: Results of a repeated cross-sectional study

**DOI:** 10.1101/2022.05.06.22274755

**Authors:** Faisal Awartani, Nouar Qutob, Mohammad Asia

## Abstract

**Background:** Seroprevalence studies provide an accurate measure of SARS-CoV-2 spread at a population level and the number of undiagnosed individuals. Repeated cross-sectional sero-studies are encouraged to monitor the spread of the virus. The aim of this study is to assess the seroprevalence rate among a random sample of Palestinians residing in the West Bank region of Palestine.

**Methods:** This second round cross-sectional study involved 1451 participants, who agreed to be interviewed and answer the questionnaire, where 910 of them agreed to participate in the sero- study and donate a blood sample to be tested for antibodies. The sample was randomly selected from the adult population, 18 years or older, living the West Bank region of Palestine.

Serological tests for 910 adequate serum samples were done using immunoassays for detection of antibodies against SARS-CoV-2. Sociodemographic information and medical history data was collected.

**Results:** Study findings indicate that as of October 2021, a seroprevalence rate of 75.9% (30% due to infection with Covid-19 virus and 45.9% due to vaccination), 95% CI (73.1-78.7). The results indicate that the prevalence of antibodies among those who are unvaccinated and undiagnosed was 45.2% with 95% CI (39.9-50.5%).

The average age of participants was 37.6 years old. 49.2% were females and 50.8% were males. In relation to COVID-19, 13.6% of respondents reported getting infected by Covid-19 with statistically significant difference (P_value=0.001) between males (10.7%) and females (16.5%).

. In terms of vaccination, 52.8% of respondents reported getting vaccinated with an important difference between males (64.3%) and females (40.9%), (P-value=0.000).

**Conclusion:** Our findings reveal a drastic rise in seroprevalence of SARS-CoV-2 antibodies due to infection and vaccination. This information is useful for assessing the degree of herd immunity among the adult population and provides better understanding of the pandemic. Population-based seroprevalence studies should be conducted periodically to monitor the SARS-CoV-2 seroprevalence in Palestine and inform policymakers about the efficacy of the surveillance system and the public compliance with vaccination policies especially among females

## INTRODUCTION

Severe acute respiratory syndrome coronavirus-2 (SARS-CoV-2), a novel virus that causes Coronavirus Disease 2019 (COVID-19) has infected over 460 million and caused the deaths of over six million people worldwide as of March 12, 2022 [1]. However, these figures underestimate the accurate cumulative prevalence or incidence of infection [2], due to lack of accessibility of diagnostic testing, limited testing capacity [3], and asymptomatic infections [4].

Globally, seroprevalence studies using serologic testing have been recognized to provide better estimates of COVID-19 infection rates and prevalence on a population level in different populations, by capturing individuals with mild or no symptoms and others who never underwent diagnostic testing [5]. Assessing the cumulative prevalence is critical to understanding disease transmission rates and for understanding the evolution of the pandemic [5, 6]. The true prevalence of infection is believed to be 12.5 times more than the number of PCR-reported infections [7] as there are many asymptomatic infections, which constitute the majority of all SARS-COV-2 infections, yet are less likely to present for diagnostic testing [8, 9]. Consequently, the accuracy of such seroprevalence studies has been doubted due to insufficient specificity of the serologic assay for a low prevalence population and due to potential sampling bias [10].

In March 2020 a nationwide lockdown was implemented in Palestine, to contain the spread of COVID-19. As of March 2022, approximately 580,165 infections have been recorded, and 5,326 people have died in Palestine [11]. Previously, the authors of this study undertook a cross-sectional study [12] which revealed 0% seroprevalence of SARS-CoV-2 among a random sample of adults in the general population in the West Bank region of Palestine. Other studies [13] conducted among the visitors of primary health care centers in Paelstine showed 36% seroprevalence. A large scael sero survey was conducted by the Paesitnian ministry of health and WHO produced a seroprevalence of 40% among the general population who are 10 years or older in the West Bank and Gaza [18]. There is a lack of data on the percentage of undiagnosed Palestinian population with previous mild or asymptomatic COVID-19. This rsecond round cross-sectional study was conducted to assess the seroprevalence of SARS-CoV-2 among a national sample of vaccinated and non-vaccinated adults in the West Bank region of Palestine, be it due to infection with COVID-19 or due to vaccination or both. A substantial seroprevalence of anti-SARS-CoV-2 antibodies in the Palestinian population should provide some measure of protection against future waves of COVID-19 in the country.

## METHODS

### Study design and participants

This second round cross-sectional study was conducted between September 14, 2021 and October 21, 2021. The study included a random sample of adults aged 18 years and above, residing in 11 governorates in the West Bank, Palestine. The sample included 1451 individuals randomly selected from households using three-stage cluster sampling. The cluster of households or census tracts is considered to be a geographic location which is comprised of 124 households. The process for conducting cluster sampling included: (1) Selection of 125 census tracts (clusters) of households using the sampling frame provided by the Palestinian Central Bureau of Statistics (PCBS), (2) Selection of 16 households randomly from each cluster and (3) selection of individuals at random from the list of individuals who are over the age of 18 and belong to the selected household. The clusters were selected using probability proportional to size (PPS) sampling, see Table (1). The following steps were used to select the number of clusters within each population location: (1) the sampling interval (eg SI=N/m) which equals the total number of households divided by the total number of clusters need to be selected by the sample was calculated, (2) a random number R0 between 0 and SI was calculated, and (3) calculated Ri as R0+i*SI, a cluster is selected in Li if Ri belongs to the interval [Ci-1, Ci].

**Table 1:** The PPS sampling algorithm for selecting population locations.

| Location | # of households in the location | Cumulative |
| --- | --- | --- |
| L1 | $X_1$ | $C_1=X_1$ |
| L2 | $X_2$ | $C_2=X_1+X_2$ |
| L3 | $X_3$ | $C_3=X_1+X_2+X_3$ |
| . | . | . |
| . | . | . |
| . | . | . |
| $L_{k-1}$ | $X_{k-1}$ | $C_{k-1}=X_1+X_2+\dots+X_{k-1}$ |
| $L_k$ | $X_k$ | $C_k = X_1+X_2+\dots+X_k$ |
| PPS, probability proportional to size. |  |  |

### Data Collection

Sociodemographic information, risk factors, medical history, and COVID-19 related information (eg. Symptoms (due to infection or due to vaccination); vaccination status) were obtained during a face-to-face interview from 1451 participants (Table 2). Upon the interview, 911 participants gave consents to test for antibodies against COVID-19. Blood samples were collected from the participants. Blood samples were centrifuged, and serum was separated, labelled and stored at -20°C at AAUP laboratory until it was used.

**Table (2):**
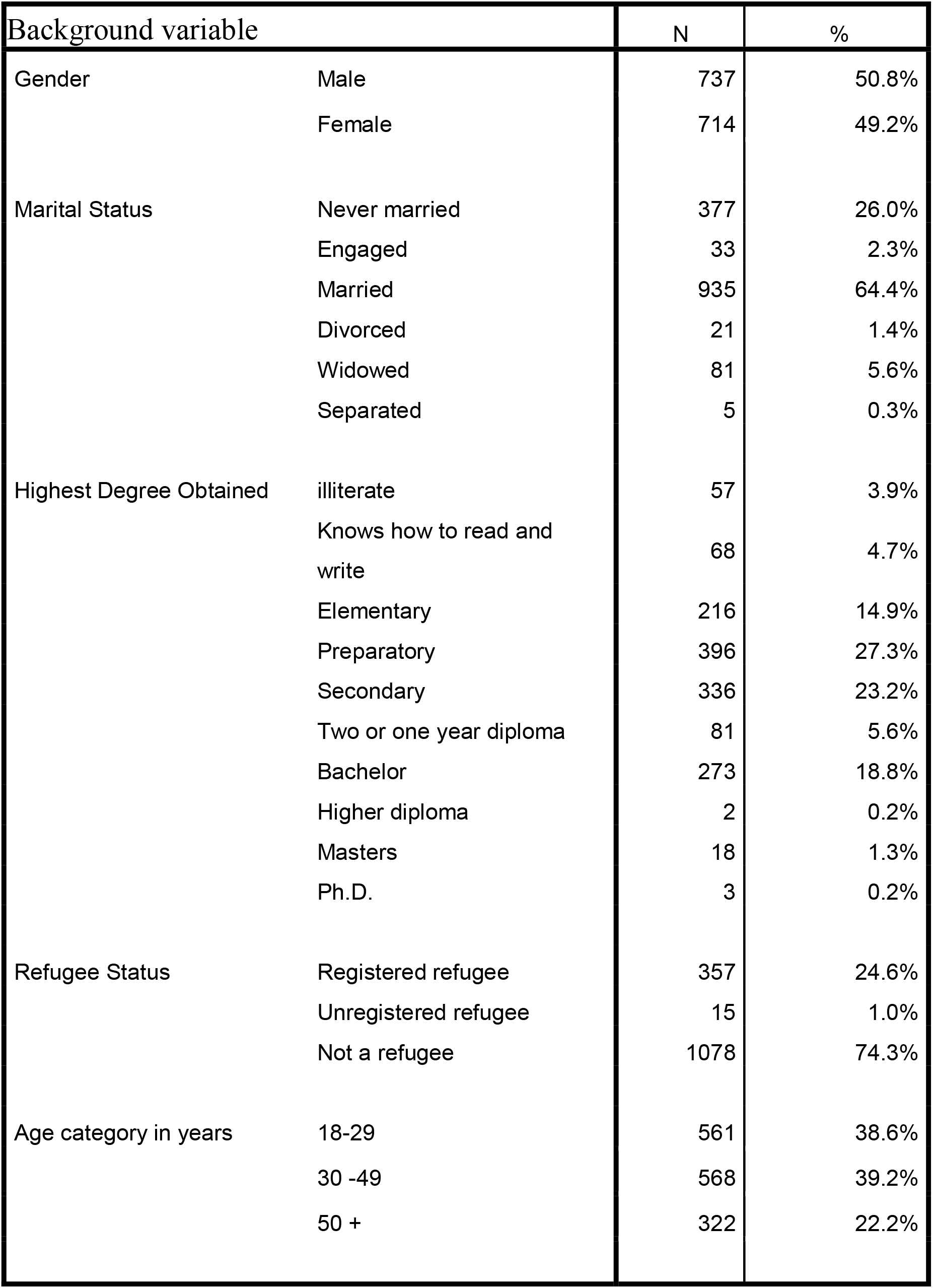
Sample Characteristics

Data collection was conducted by healthcare professionals following standardized health protocols (World Health, 2020). Ethics approval was obtained from the Helsinki committee ((PHRC/HC/737/20).) Participants recruited in the study gave written consent forms prior to participation in the study voluntary agreeing to participate in the research.

The study was conducted between September 14^th^ 2021 and October 21^st^. Demographic and health Data was collected through face to face interviews with the 1451 participants. Blood samples were drawn from 911 participants who agreed to participate in the sero study.

### Sars-CoV-2 Antibody Testing

Serological tests for the 911 adequate serum samples were done using an Elecsys^®^ Anti-SARS-CoV-2 assay by using the Cobas Analyzer cobas e 411 (Roche). The assay is an immunoassay that uses a recombinant protein representing the nucleocapsid (N) antigen. Serum samples in which the nucleocapsid (N) antigen was not detected were run using Elecsys Anti-SARS-CoV-2 S by using the Cobas Analyzer cobas e 411 (Roche). The assay detects antibodies to SARS□CoV□2 spike protein RBD which are produced by mRNA vaccines. Samples from recovered and vaccinated cases with detected SARS-CoV-2 anti N and SARS-CoV-2 anti S antibodies were run as positive controls.

### Statistical Analysis

Both univariate and bivariate inferential statistical methods were used for statistical analysis. Confidence interval using 95% confidence levels are used for estimation the seroprevalence across different segments within the Palestinian adult population living in the West Bank region. The lower and upper confidence limits of were calculated. Cross-tabulation method was used to explore bivariate relationships between seroprevalence and gender among other background variables. Chi-square test was utilized for testing if the statistical significance of bivariate relations among the different variables.

## RESULTS

### Sample Characteristics

In total, 1451 participants were enrolled in the study in which 911 undertook serological testing and blood sampling. Among these, 50.8% were males, 64.4% were married, 27.3% finished preparatory school, and 74.4% were non-refugees. The mean age of participants is 37.6. See Table (2) for detailed sample characteristics.

The main findings of the survey showed that a large majority of adult Palestinians (75.9%) developed antibodies against COVID-19. The results did not show significant difference (P-value=0.23) between males (74.2%) and females (77.5%) in the overall seroprevalence, see Table (3). While there was a strong difference according to gender when we accounted for the source of antibodies whether it was due to infection or vaccination. The results showed that the prevalence of antibodies due to infection among females was twice (41.2%) that among males (19.9%), (P-value=0.000). See Table (6).

**Table (3):**
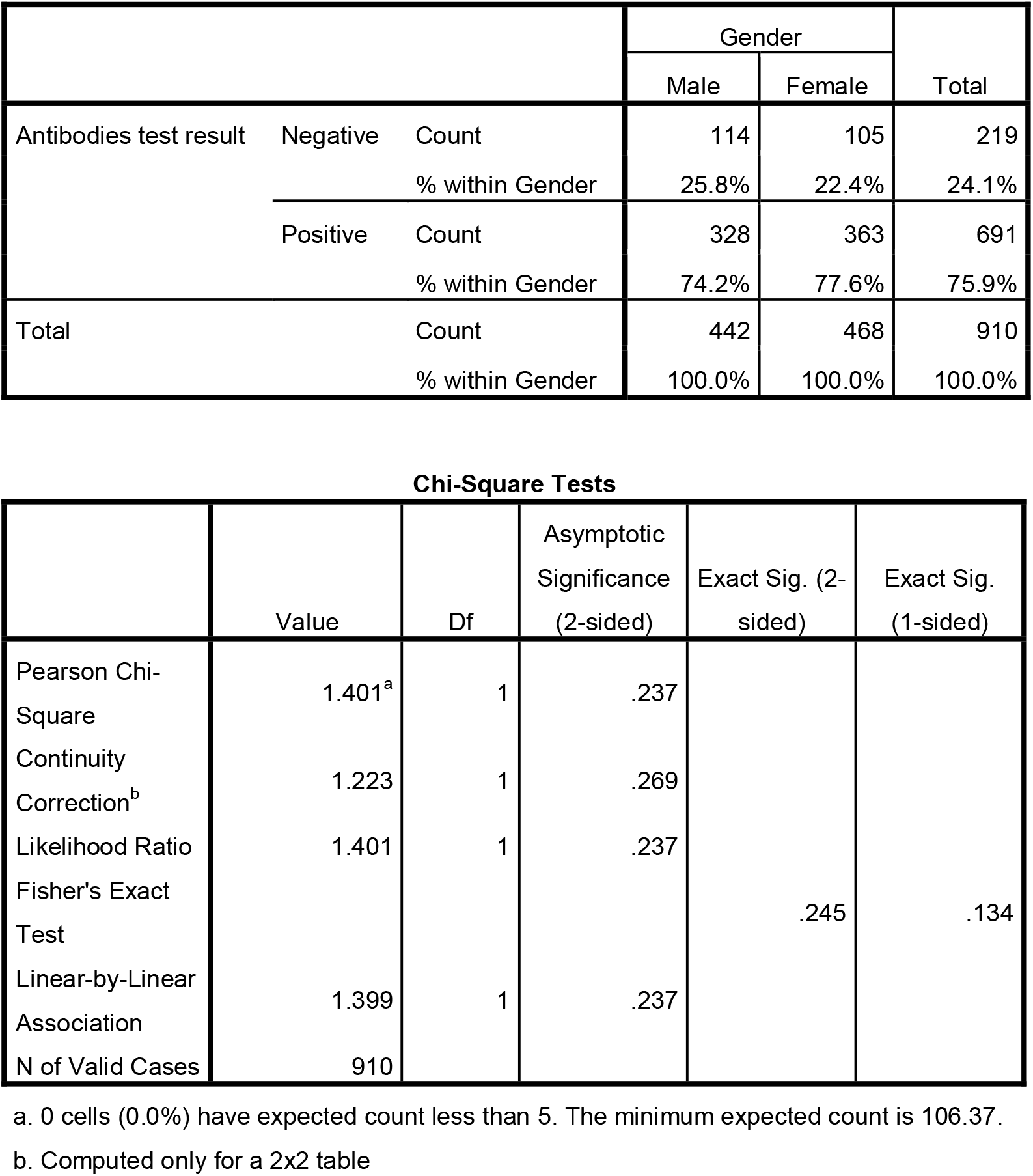
Antibodies test results irrespective of he source (infection or vaccine) by gender

**Table (4):**
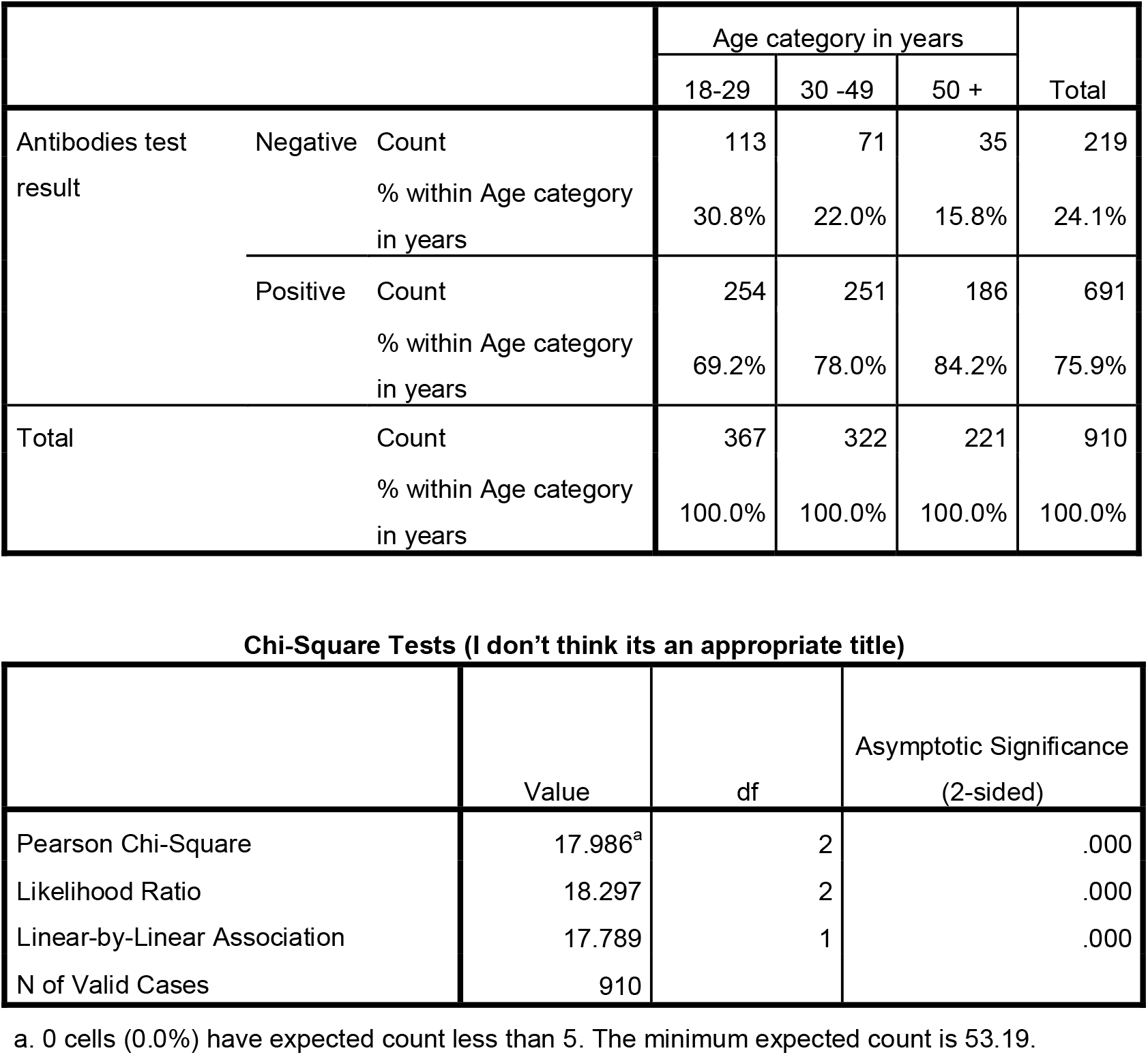
Antibodies test results irrespective of the source (infection or vaccine) by age category

**Table (5):** Antibodies test results by vaccination and Covid-19 infection status as reported by respondents

|  |  |  | Antibodies test result |  | Total |
| --- | --- | --- | --- | --- | --- |
|  |  |  | Negative | Positive |  |
| Vaccination and Covid-19 infection status as reported by the respondents | Got infected with Covid-19 and got vaccinated | Count | 1<br>1.7% | 59<br>98.3% | 60<br>100.0% |
|  | Got infected with Covid-19 and did not get vaccinated | Count | 4<br>5.6% | 67<br>94.4% | 71<br>100.0% |
|  | Did not get infected with Covid-19 and got vaccinated | Count | 26<br>6.0% | 410<br>94.0% | 436<br>100.0% |
|  | Did not get infected with Covid-19 and did not get vaccinated. | Count | 188<br>54.8% | 155<br>45.2% | 343<br>100.0% |
| Total |  | Count | 219<br>24.1% | 691<br>75.9% | 910<br>100.0% |

Chi-Square Tests
|  | Value | Df | Asymptotic Significance (2-sided) |
| --- | --- | --- | --- |
| Pearson Chi-Square | 285.275<sup>a</sup> | 3 | .000 |
| Likelihood Ratio | 294.040 | 3 | .000 |
| Linear-by-Linear Association | 62.356 | 1 | .000 |
| N of Valid Cases | 910 |  |  |

a. 0 cells (0.0%) have expected count less than 5. The minimum expected count is 14.44.

**Table (6):**
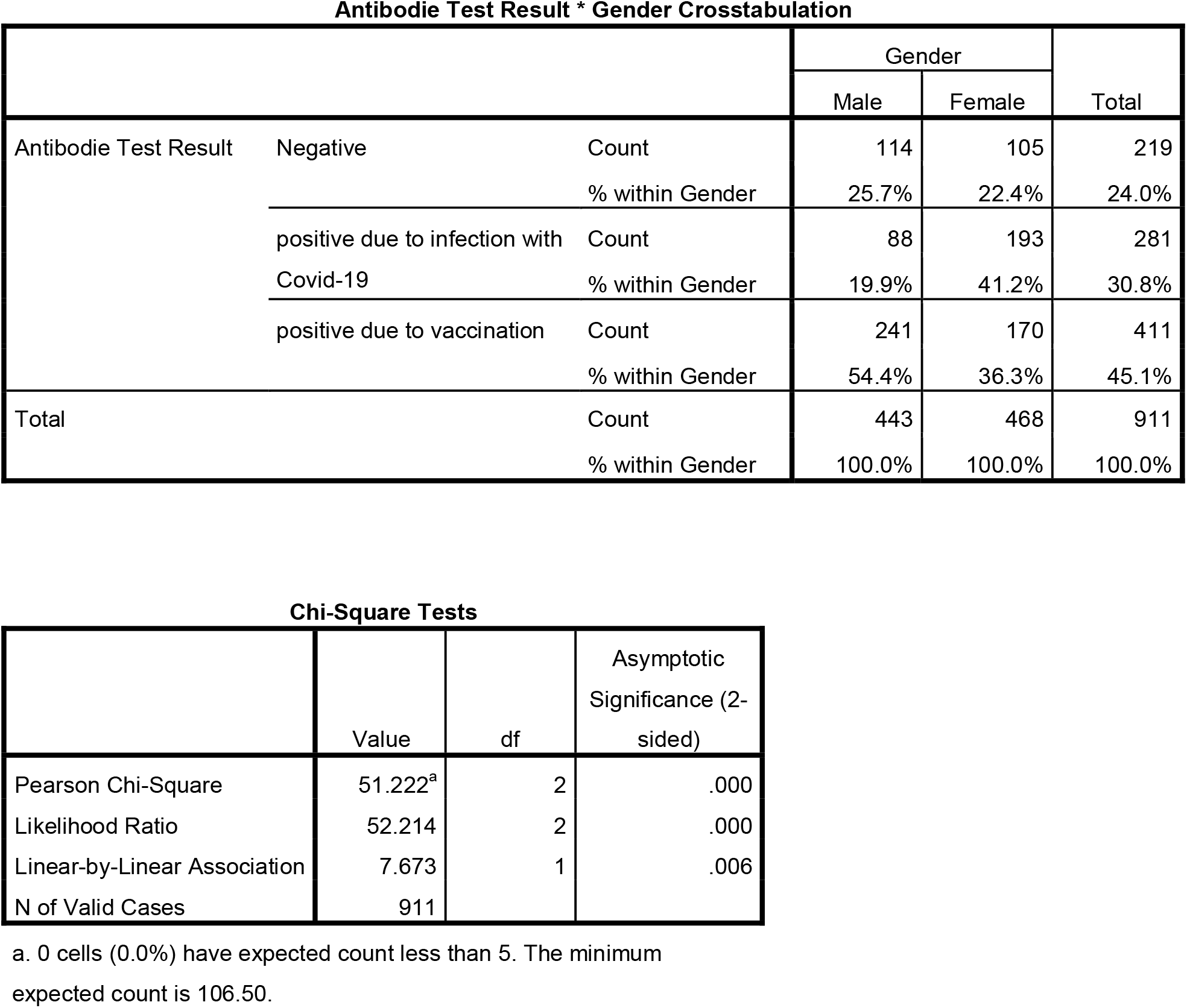
Seroprevalence according to gender and source of antibodies (infection or vaccine)

The study results showed quite a significant difference (P-value=0.000) in antibodies prevalence across age groups in the Palestinians society. The youngest age group (18-29 years) recorded the lowest prevalence of antibodies (69.2%), while the age group (30 -50 years) showed a prevalence of 77.9%. While the older group (50 years or older) got the highest prevalence of antibodies (84.1%). Such result could be attributed to the fact that the older population categories got more vaccinated than the younger population categories, see Table (4).

An important factor that explained the variation in levels of antibodies prevalence is the vaccination and infection status among the Palestinian population. A new variable was computed based on the infection status with Covid-19 and the vaccination status. The new variable was used to segment the Palestinian population over the age of 18 into four categories described as follows: a) individuals who got infected and got vaccinated, b) individuals who got infected and did not get vaccinated, c) individuals who did not get infected and got vaccinated, d) individuals who did not get infected and did not get vaccinated.

The largest group of individuals within the Palestinian adult population living in the West Bank region are those who did not get infected and got vaccinated (46.4%) followed by the second largest group, which represents those who did not get vaccinated and reported (thought) that they did not get infected (40.0%), see Figure (1).

**Figure (1):**
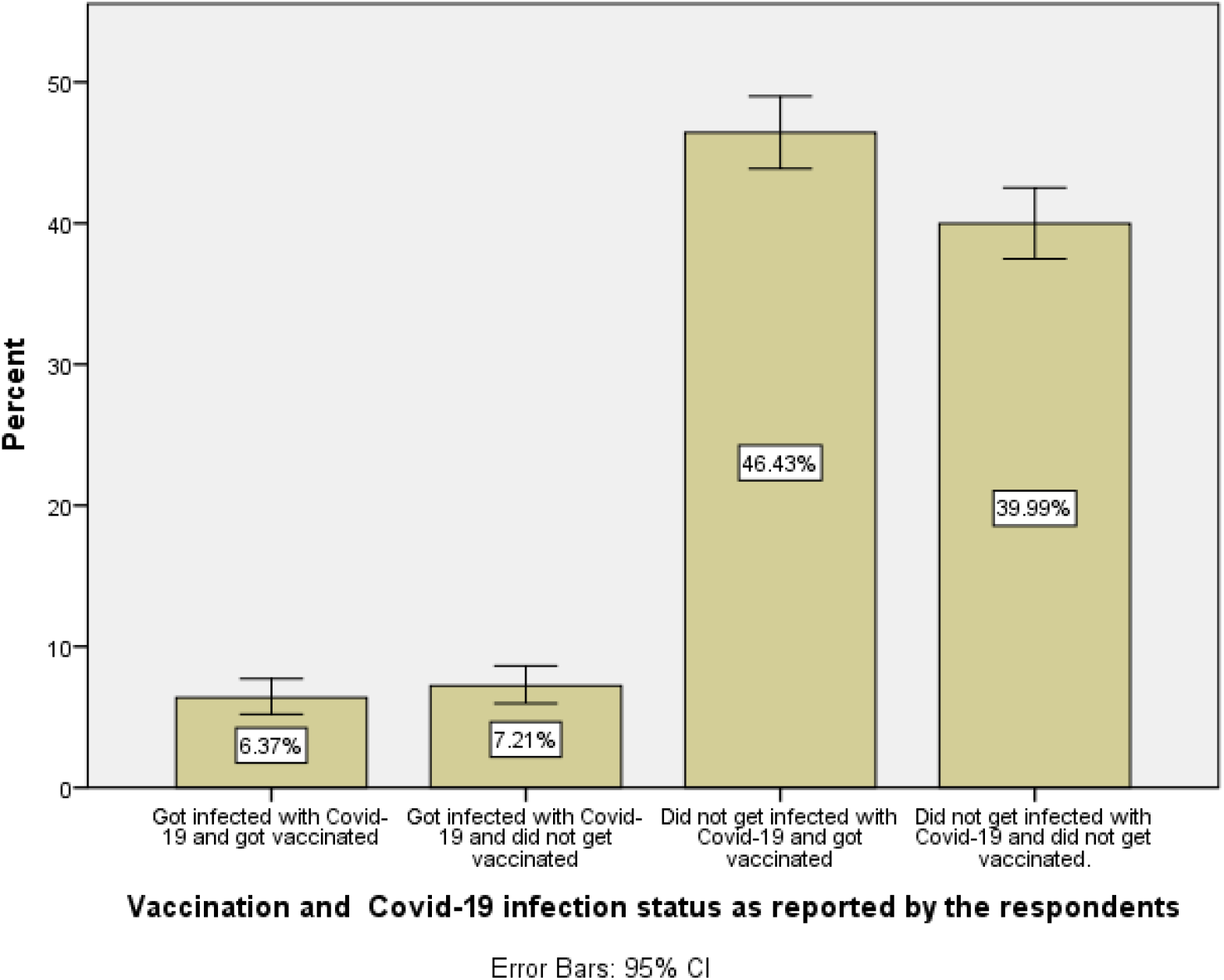
Distribution of vaccination and Covid-19 infection status for adult Palestinians living in the West Bank region of Palestine.

Our findings showed a strong bivariate relationship between antibody test results and infection-vaccination status (P-value=0.000), see Table (5). It is evident, the antibodies prevalence among those who did not get infected and got vaccinated is 94.0% in comparison to 45.2% among those who reported that they did not get vaccinated and did not get infected as well. Interestingly, 45.2% of those who did not get vaccinated and reported that they did not get infected with Covid-19 actually got infected without knowing, concluding this group of the population were asymptomatic to the infection.

## DISCUSSION

The present study reports the seroprevalence of anti-SARS-CoV-2 antibodies as of October 2021. There have not been many studies assessing seroprevalence after the beginning of vaccinations. According to this study, seroprevalence of antibodies due to infection with COVID-19 or vaccination one and half years after the start of the pandemic and several□months after the vaccination rollout was 75.9%, varying among age groups, with the youngest age group having the highest seroprevalence. The high seroprevalence □rate of antibodies observed in our cohort was surely influenced by the high rate of vaccination within the Palestinian adults living in the West Bank region. This finding was similar to a study conducted in Chile in which reported a similar seroprevalence of 77.4% five months after initiation of the vaccination [14]. Similar to the Chile study, the results of our study showed a significant difference across gender in antibodies prevalence when we account for the source of positivity with antibodies whether it’s due to infection with Covid-19 or due to vaccination, see Table (6). This result is consistent with other studies which found a statistically significant difference between prevalence of antibodies and gender, [14],[15]. At the regional level, a three phased seroprevalence survey was conducted in Jordan, where results revealed that seroprevalence dramatically increased over time, with only a tiny fraction of seropositive individuals in August 2020 (0.3%), to increase up to more than 20- fold in October 2020 (7.0%) and to reach one-third of the overall population exposed by the end of 2020 (34.2%), [15]. Such results are compatible with similar seroprevalence survey conducted by the Palestinian ministry of health and WHO at the end of 2020 [18]. While similar results were obtained by a seroprevalence survey conducted by Arab American university in during June 2020. [12]. The most recent serology survey conducted by CDC over the period of September 2021 till December 2021 in the US, Seroprevalence increased from 36.5% (95% CI = 35.7–37.4) to 63.7% (95% CI = 62.5–64.8) among adults aged 18–49 years, 28.8% (95% CI = 27.9–29.8) to 49.8% (95% CI = 48.5–51.3) among those aged 50–64 years, and from 19.1% (95% CI = 18.4–19.8) to 33.2% (95% CI = 32.2–34.3) among those aged ≥65 years, [16]. Approximately similar seroprevalence due to infection with Covid-19 (30%) was obtained by our serology survey that targeted adults over the age of 18 year during September 2021. See Table (6).

Two years after the start of the COVID-19 pandemic, the actual prevalence of infection is still underestimated compared with PCR-confirmed COVID-19 cases. Older compared with younger individuals have lower antibody levels after vaccination. Approximately, 40% of the Palestinian adult population over the age of 18 years thought they were not infected and did not make the effort to get vaccinated. The study’s findings showed 45% of those who thought they were not infected and did not get vaccinated, tested positive for SARS Covid-2 antibodies. By multiplying these two percentages, one can conclude that 18% of the Palestinian adult population who are over the age 18 got infected with SARS Covid-2 without knowing. This group of people could have contributed to a faster transmission rate in the spread of the virus, since they were asymptomatic. Governments and public health officials should put more effort to detect asymptomatic cases through conducting seroprevalence surveys on a regular basis and develop regulations to encourage more people to get vaccinated. Tracking seroprevalence due to vaccination is an important issue, since vaccination contributes to the reduction of hospitalization among Covid-19 patients. A preliminary evidence has highlighted a possible association between severe COVID-19 and persistent cognitive deficits[17]. Further research is required to confirm this association, determine whether cognitive deficits relate to clinical features from the acute phase or to mental health status at the point of assessment, and quantify rate of recovery.

## Data Availability

All data produced in the present study are available upon reasonable request to the authors

## Acknowledgements

The authors would like to thank PCBS staff who participated and managed data collection process. We would like to thank AAUP nursing school for the work they did on blood sample collection from participants. Finally, we would like to thank AAUP lab technicians and manager for their hard work on the blood samples analysis.

## Contributors

Original Draft—Faisal Awartani. Writing Editing and Reviewing—Faisal Awartani, Nouar Qutub, Mohammed Asia. Lab Tests—Nouar Qutob. Data Analysis and Visualization--Faisal Awartani.

## Funding

The authors have not declared any specific grant from any funding agency in the public, commercial or non-for-profit sector.

## Competing Interests

None Declared.

## Patient consent for publication

Not required.

## Notes

### Competing Interest Statement

The authors have declared no competing interest.

### Funding Statement

This study was funded by Arab American University of Palestine (www.aaup.edu)

### Author Declarations

Data collection was conducted by healthcare professionals at Arab American University of Palestine following standardized health protocols (World Health, 2020). Approval from the National Ethical Committee was obtained (PHRC/HC/737/20). Written informed consent was obtained from all participants before the interview and the blood sampling and approval of utilization of tests was obtained from Medicare laboratories Participation was completely voluntary and all participants were informed that all the information collected would be anonymous and treated as confidential.

## REFERENCE

1 World Health Organization. WHO coronavirus (COVID-19) dashboard. Geneva: World Health Organization; 2021. https://covid19.who.int/table (accessed 12 Mar 2022).

2 Cheng MP, Papenburg J, Desjardins M, et al. Diagnostic Testing for Severe Acute Respiratory Syndrome-Related Coronavirus 2: A Narrative Review. Ann Intern Med. 2020;172(11):726–34.

3 Lieberman-Cribbin W, Tuminello S, Flores RM, et al. Disparities in COVID-19 Testing and Positivity in New York City. Am J Prev Med. 2020;59(3):326–32.

4 Buitrago-Garcia D, Egli-Gany D, Counotte MJ, et al. Occurrence and transmission potential of asymptomatic and presymptomatic SARS-CoV-2 infections: A living systematic review and meta-analysis. PLoS Med. 2020;17(9):e1003346.

5 Busch MP, Stone M. Serosurveillance for Severe Acute Respiratory Syndrome Coronavirus 2 (SARS-CoV-2) Incidence Using Global Blood Donor Populations. Clin Infect Dis. 2021;72(2):254–6.

6 Havers FP, Reed C, Lim T, et al. Seroprevalence of Antibodies to SARS-CoV-2 in 10 Sites in the United States, March 23-May 12, 2020. JAMA Intern Med. 2020.

7 Bajema KL, Wiegand RE, Cuffe K, et al. Estimated SARS-CoV-2 Seroprevalence in the US as of September 2020. JAMA Intern Med. 2021;181(4):450–60.

8 Huang L, Zhang X, Zhang X, et al. Rapid asymptomatic transmission of COVID-19 during the incubation period demonstrating strong infectivity in a cluster of youngsters aged 16-23 years outside Wuhan and characteristics of young patients with COVID-19: A prospective contact-tracing study. J Infect. 2020;80(6):e1–e13.

9 McLaughlin CC, Doll MK, Morrison KT, et al. High Community SARS-CoV-2 Antibody Seroprevalence in a Ski Resort Community, Blaine County, Idaho, US. Preliminary Results. medRxiv. 2020:2020.07.19.20157198.

10 Bobrovitz N, Arora RK, Cao C, Boucher E, Liu M, Donnici C, et al. Global seroprevalence of SARS-CoV-2 antibodies: A systematic review and meta-analysis. PLoS One. 2021;16(6):e0252617.

11 Worldometer. State of Palestine: Coronovirus cases. 2022. https://www.worldometers.info/coronavirus/country/state-of-palestine (accessed 17 Mar 2022).

12 Qutob N, Awartani F, Salah Z, et al. Seroprevalence of SARS-CoV-2 in Palestine: a cross-sectional seroepidemiological study. medRxiv. 2020:2020.08.28.20180083.

13 Maraqa B, Basha W, Khayyat R, et al. Prevalence of SARS-CoV-2 antibodies in the Palestinian population: A primary health center-based cross-sectional study. PLos One. 2021;16(10):e0258255.

14 Sauré D, O’Ryan M, Torres JP, et al. Dynamic IgG seropositivity after rollout of CoronaVac and BNT162b2 COVID-19 vaccines in Chile: a sentinel surveillance study. The Lancet Infectious Diseases. 2022;22(1):56–63

15 Bellizzi S, Alsawalha L, Sheikh Ali S, Sharkas G, Muthu N, Ghazo M, Hayajneh W, Profili MC, Obeidat NM. A three-phase population based sero-epidemiological study: Assessing the trend in prevalence of SARS-CoV-2 during COVID-19 pandemic in Jordan. One Health. 2021 Jul 10;13:100292. doi: 10.1016/j.onehlt.2021.100292. PMID: 34295958; PMCID: PMC8272624.

16 Clarke KE, Jones JM, Deng Y, et al. Seroprevalence of Infection-Induced SARS-CoV-2 Antibodies — United States, September 2021–February 2022. MMWR Morb Mortal Wkly Rep 2022;71:606–608. DOI: http://dx.doi.org/10.15585/mmwr.mm7117e3ext

17 Adam Hampshire, Doris A. Chatfield, Anne Manktelow MPhil, Amy Jolly, William Trender, Peter J. Hellyer, Martina Del Giovane, Virginia F.J. Newcombe, Joanne G. Outtrim, Ben Warne, Junaid Bhatti, Linda Pointon, Anne Elmer, Nyarie Sithole, John Bradley, Nathalie Kingston, Stephen J. Sawcer, Edward T. Bullmore, James B. Rowe, David K. Menon, Multivariate profile and acute-phase correlates of cognitive deficits in a COVID-19 hospitalised cohort, eClinicalMedicine, Volume 47, 2022, 101417, ISSN 2589-5370,

18 Seroprevalence of COVID-19 in Palestine in 2020 I Rayan, SE Qaddomi, O Najjar, S Abbas, K Mousa, L Iraqi, E Abdelkreem Aly, K Abu Khader, A Barakat, R Salman medRxiv 2021.10.04.21263131; doi: https://doi.org/10.1101/2021.10.04.21263131

